# Research on Artificial Intelligence and Primary Care: A Scoping Review

**DOI:** 10.1101/19003913

**Authors:** Jacqueline K. Kueper, Amanda L. Terry, Merrick Zwarenstein, Daniel J. Lizotte

## Abstract

**Objective:** The purpose of this study was to assess the nature and extent of the body of research on artificial intelligence (AI) and primary care.

**Methods:** We performed a scoping review, searching 11 published and grey literature databases with subject headings and key words pertaining to the concepts of 1) AI and 2) primary care: MEDLINE, EMBASE, Cinahl, Cochrane Library, Web of Science, Scopus, IEEE Xplore, ACM Digital Library, MathSciNet, AAAI, arXiv. Screening included title and abstract and then full text stages. Final inclusion criteria: 1) research study of any design, 2) developed or used AI, 3) used primary care data and/or study conducted in a primary care setting and/or explicit mention of study applicability to primary care; exclusion criteria: 1) narrative, editorial, or textbook chapter, 2) not applicable to primary care population or settings, 3) full text inaccessible in the English Language. We extracted and summarized seven key characteristics of included studies: overall study purpose(s), author appointments, primary care functions, author intended target end user(s), target health condition(s), location of data source(s) (if any), subfield(s) of AI.

**Results:** Of 5,515 non-duplicate documents, 405 met our eligibility criteria. The body of literature is primarily focused on creating novel AI methods or modifying existing AI methods to support physician diagnostic or treatment recommendations, for chronic conditions, using data from higher income countries. Meaningfully more studies had at least one author with a technology, engineering, or math appointment than with a primary care appointment (57 (14%) compared to 217 (54%)). Predominant AI subfields were supervised machine learning and expert systems.

**Discussion:** Overall, AI research associated with primary care is at an early stage of maturity with respect to widespread implementation in practice settings. For the field to progress, more interdisciplinary research teams with end-user engagement and evaluation studies are needed.

**SUMMARY BOXES:** *Section 1: What is already known on this topic:*

- Advancements in technology and the availability of health data have increased opportunities for artificial intelligence to be used for primary care purposes.
- No comprehensive review of research on artificial intelligence associated with primary care has been performed.

*Section 2: What this study adds:*

- The body of research on artificial intelligence and primary care is driven by authors without appointments in primary care departments and is focused on developing artificial intelligence methods to support diagnostic and treatment decisions.
- There is a need for more interdisciplinary research teams and evaluation of artificial intelligence projects in ‘real world’ practice settings.

## INTRODUCTION

Artificial Intelligence (AI) has gained prominence through widespread, high-profile media, academic, and industry initiatives (1–6). AI research began in the 1950s, but the recognition of its potential for adoption in health care settings is widespread and ongoing (7–9). Applications of AI to the foundational sector of health care, Primary Care (PC), are particularly compelling and promoted by advances in technology and the availability of data such as through increased use of Electronic Medical Records (EMRs) (10–12).

From its inception in the 1950s, AI was primarily concerned with processes by which computers might achieve ‘intelligence’ comparable to that of humans, and how we might recognize such intelligence (13). Turing’s (1950) seminal paper, “Computing Machinery and Intelligence,” was concerned more with the latter, but the work sparked a rich diversity of research activities (13,14). The field of AI now encompasses a wide variety of methodology, much of which falls into two broad categories: rule-centred and data-centred. Rule-centred methods came from the study of logical reasoning, and are intended to capture intelligence by explicitly writing down the rules that govern it and then deploying that intelligence to carry out different tasks (13). Data-centric methods like machine learning have focused more on learning to perform specific tasks using previously collected data rather than explicitly provided rules (13). Definitions of select AI subfields with examples are provided in Table 1S (Appendix C) and example health applications are presented below.

The first rule-centred AI application for health care, MYCIN, was not in PC. MYCIN is an expert system developed in the 1970s to diagnose blood infections using over 450 rules derived from experts, textbooks, and case reports (13,15). Although met with initial excitement, over time, limitations were identified in rule-centric methods in terms of handling the complexity of real-world decisions. At the same time, increasing amounts of health data promoted a shift towards data-centric methods in machine learning, which were thought to be better able to capture complex relationships between health-relevant features from data than could be represented using rules. Machine learning methods are now used to reveal patterns in data to answer health research questions, such as to facilitate prediction of diabetes or cancer diagnoses (16–19). Machine learning methods figure prominently in natural language processing, a sub-field of AI, which can be applied to unstructured text data to extract structured information or summarize documents (20). The use of AI to input or process EMR data into formats for future use or to output meaningful summary information could potentially remove some of the EMR-associated burden from clinicians (21–23). Other sources of health data are also processed by AI methods; for example, skin cancer diagnosis using computer vision and machine learning has advanced due to 1) skin lesion imaging data being readily available with easy digitization and image recognition technology, and 2) the suitability of using supervised machine learning for predicting the probability that a lesion is pathological (24,25). These types of techniques are purported to diagnose at least as accurately as clinicians; however, the data are mainly from settings past the point of referral from PC, where the distribution and spectrum of lesions is wider, and clinicians are restricted to visual inspection—further evidence is required to assess usefulness of these tools for PC settings (24,25). Several commentaries and reports describe these and other applications of AI to health, though not predominantly for PC (1,2,5,6,17,23,26,27).

Barbara Starfield (1998) defines PC as the “level of a health service system that provides entry into the system for all new needs and problems, provides person-focused (as opposed to disease-oriented) care over time, provides care for all but very uncommon or unusual conditions, and coordinates or integrates care provided elsewhere or by others” (10). AI may increase the range of health conditions that can be handled in PC or support care providers with information retrieval and focusing on aspects of PC like relationship building and shared decision making (9,23,26). However, there is no comprehensive review of what contribution it has made so far, and thus little guidance on how best to proceed with AI research in PC.

AI’s immediate usefulness is not guaranteed: EMRs were predicted to transform PC for the better, but led to unanticipated outcomes and barriers to adoption (11,28–30). Similarly, AI could benefit PC but also do harm. Careless or ill-informed development or use of AI may exaggerate racial, class, or gender biases if models are built with biased data or used with new populations for whom performance may be poor; liability, trust, and disrupted workflow are further concerns to be addressed (9).

Optimism surrounding the use of AI to benefit patients, care providers, and society exists, yet we have a lack of knowledge about AI for PC. Therefore, we set out to locate and summarize all research literature regarding AI and PC. The objective of this study was to assess the nature and extent of the body of research on AI for PC purposes.

## METHODS

We performed a scoping review according to guidelines from Arksey (2005), Levac (2010), and Tricco (2016) whereby all literature on a topic is located through a systematic search strategy, data are extracted from each relevant study, and then findings are synthesized (31–33). We followed the PRISMA-ScR Checklist (34) (Appendix A). Our protocol was registered with the Open Science Framework on April 6, 2018 before completing our database searches (osf.io/w3n2b). There was no patient or public involvement in our study.

### Search Strategy

Search strategies were developed in collaboration with a medical sciences librarian. An initial search was run in PubMed using Medical Subject Headings for AI and PC to locate relevant documents, whereby subject headings and keywords from these documents were used to develop comprehensive search strategies in conjunction with topic area knowledge and discussion amongst DJL, ALT, and JKK. Final search strategies developed for health sciences, computer science, and interdisciplinary databases included keywords and, where possible, subject headings pertaining to the concepts of 1) AI and 2) PC. Due to subject-area terminology differences, certain terms were used exclusively in health sciences or in computer science databases (e.g. “knowledge base”). Search strategies were refined in an iterative fashion to balance comprehensiveness with feasibility; all relevant documents from the initial search were re-identified using a final search strategy in MEDLINE. Appendix B contains final search strategies for the 11 published or grey literature databases that were searched with retrieved references uploaded into Covidence (35) and duplicate documents removed: MEDLINE, EMBASE, Cinahl, Cochrane Library, Web of Science, Scopus, IEEE Xplore, ACM Digital Library, MathSciNet, AAAI, arXiv. Where possible, English-language limits were set; to get a sense of the amount of literature potentially missed, searches were re-run for a subset of the databases (MEDLINE-Ovid, CINAHL, Web of Science) with language limits re-set to accept all languages except English. Fewer than 10 documents were retrieved from each search.

### Study Selection

#### Level 1

Titles and abstracts were independently assessed by two reviewers (JKK, DJL) according to eligibility criteria: 1) identify the document as reporting on a research study, 2) mention or allude to the use of AI, and 3) mention a PC data source, setting, or type of personnel (e.g. general practitioner, nurse). Documents were independently rated as ‘yes’ or ‘no’ against these inclusion criteria requiring two ‘yes’ votes to be included in Level 2 screening. An initial pilot test screening of the first 25 documents was performed, during which disagreements were discussed to ensure a mutual understanding of the screening criteria and to ensure capturing of relevant literature. A second, similar meeting was held after screening an additional 100 records. Disagreements from the remainder of Level 1 screening were resolved by a third reviewer (ALT).

During this process we found a substantial amount of literature on computerized cognitive behavioural therapy. For example, computer-based programs that patients can access outside of a clinic to be guided through therapy sessions (36–40). Because the methods driving these tools were often unclear and comprehensive reviews on these systems have been performed previously, these studies were excluded (n=37).

#### Level 2

Two reviewers (JKK, DJL) independently reviewed the full text of all documents that made it through Level 1 screening according to the following eligibility criteria. Inclusion criteria: 1) research study of any design, 2) developed or used AI, 3) used PC data and/or study conducted in a PC setting and/or explicit mention of study applicability to PC; exclusion criteria: 1) narrative, editorial, or textbook chapter, 2) not applicable to PC population or settings, 3) full text inaccessible in the English Language. Similar to Level 1 screening, pilot testing and refinement of eligibility criteria was performed. All disagreements were resolved by discussion until consensus (JKK, DJL).

A notable challenge that arose in assessing eligibility criteria was authors’ use of terminology that overlaps with AI when the methods used are not considered AI. For example, “data mining” or “natural language processing” was used to describe methodologies like simple database queries or string matching that are not AI. We also excluded 34 studies because there was not enough information to determine whether AI was involved, even after consulting any methods references.

### Data Extraction and Synthesis

An initial data extraction form was discussed amongst all reviewers (JKK, ALT, MZ, DJL) after pilot testing extraction of three randomly selected articles (41–43). A revised data extraction sheet was pilot tested by three reviewers (JKK, ALT, DJL) with five new randomly selected articles (25,44–47). Extractions were compared and discussed to ensure all relevant information could be captured in a consistent manner. Remaining documents were split alphabetically amongst three reviewers to be extracted independently (100 by ALT, 50 by DJL, 250 by JKK). The final version of the data extraction sheet collected information about publication details, study purpose(s), author appointment(s), PC function(s), author intended target end user(s), target health condition(s), location of data source(s) (if any), utilized subfield(s) of AI, the reviewer who extracted the data, and any reviewer notes. We agreed upon definitions for each data extraction field (Table 1S, Appendix C). For fields except author appointments and additional notes, we predefined categories based on the pilot testing and on content knowledge—studies were recorded as belonging to as many categories as were relevant. An “Other” field was used to capture specifics of studies that did not fit into a predefined category and an “Unknown” category was used if not enough information was provided for category selection. Results were summarized as categorical variables for the seven data extraction fields; meaningful cross tabulations and additional summary counts across categories were performed.

**Table 1:**
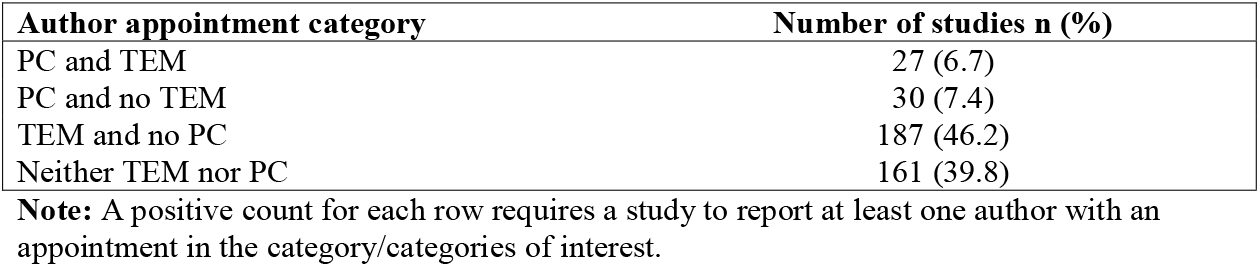
Author Appointments.

## RESULTS

### Searches

Not including duplicates our search retrieved 5,515 documents for title and abstract screening. Of those, 727 met the eligibility criteria for full text screening and 405 met the final inclusion criteria (Figure 1; a reference list for included studies is in Appendix D). The AI and PC study with the earliest date of publication, 1986, developed a supervised machine learning method to assist clinicians with abdominal pain diagnoses (48). Characteristics of all studies are summarized below according to the seven data extraction categories, which are defined in Table 1S (Appendix C).

**Figure 1:**
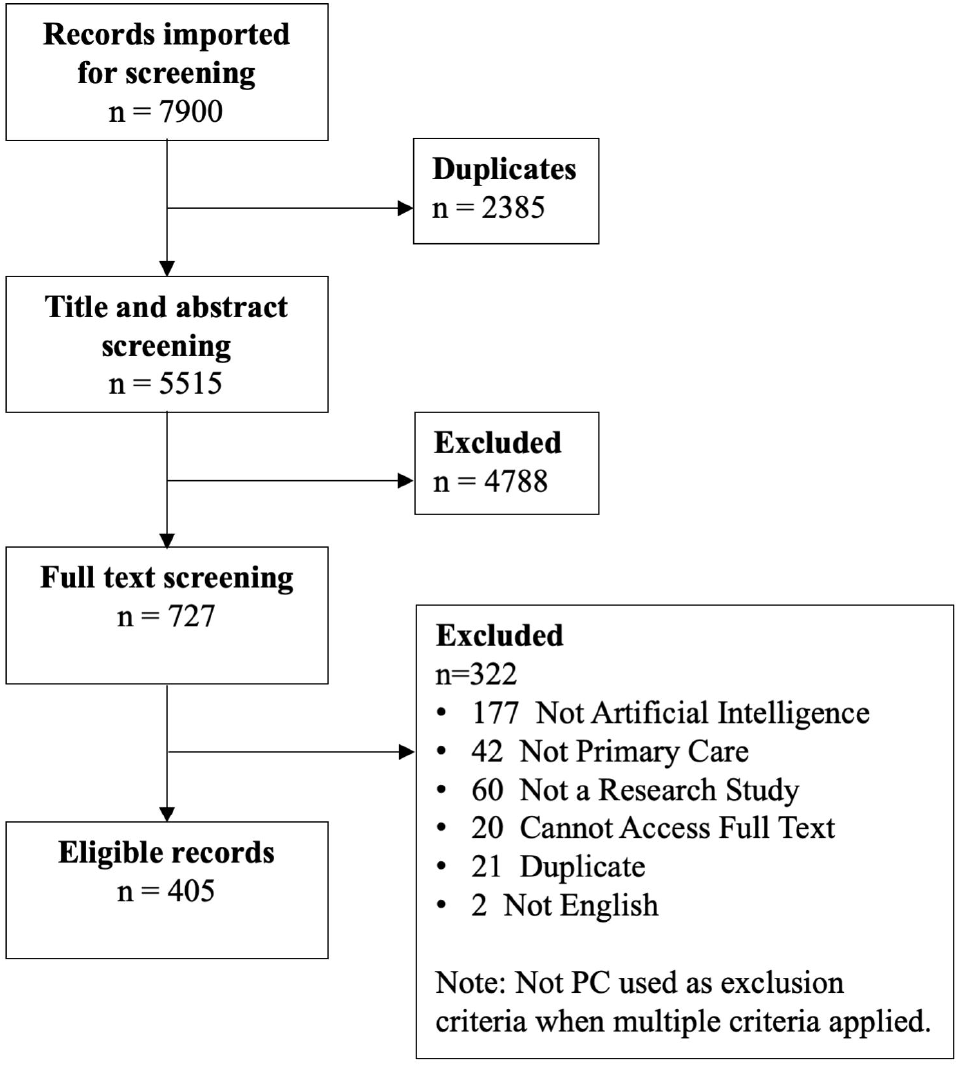
PRISMA flow diagram.

### Study Purpose

The majority of studies (n=270, 66.7%) focused primarily on developing new or adapting existing AI methods using secondary data. The second most common study purpose (n=86, 21.2%) was analysing data using AI techniques, such as eliciting patterns from health data to facilitate research (e.g. to improve scheduling or cost management). Few studies evaluated AI in a ‘real world’ setting (n=28, 6.9%).

There were series of studies that reported on multiple stages of a project, from AI development to pilot testing; these studies included intended end users located in a PC setting (49–56). A minority of studies (n=21, 5.2%) had more than one study purpose. Figure 2 presents all combinations of these studies.

**Figure 2.**
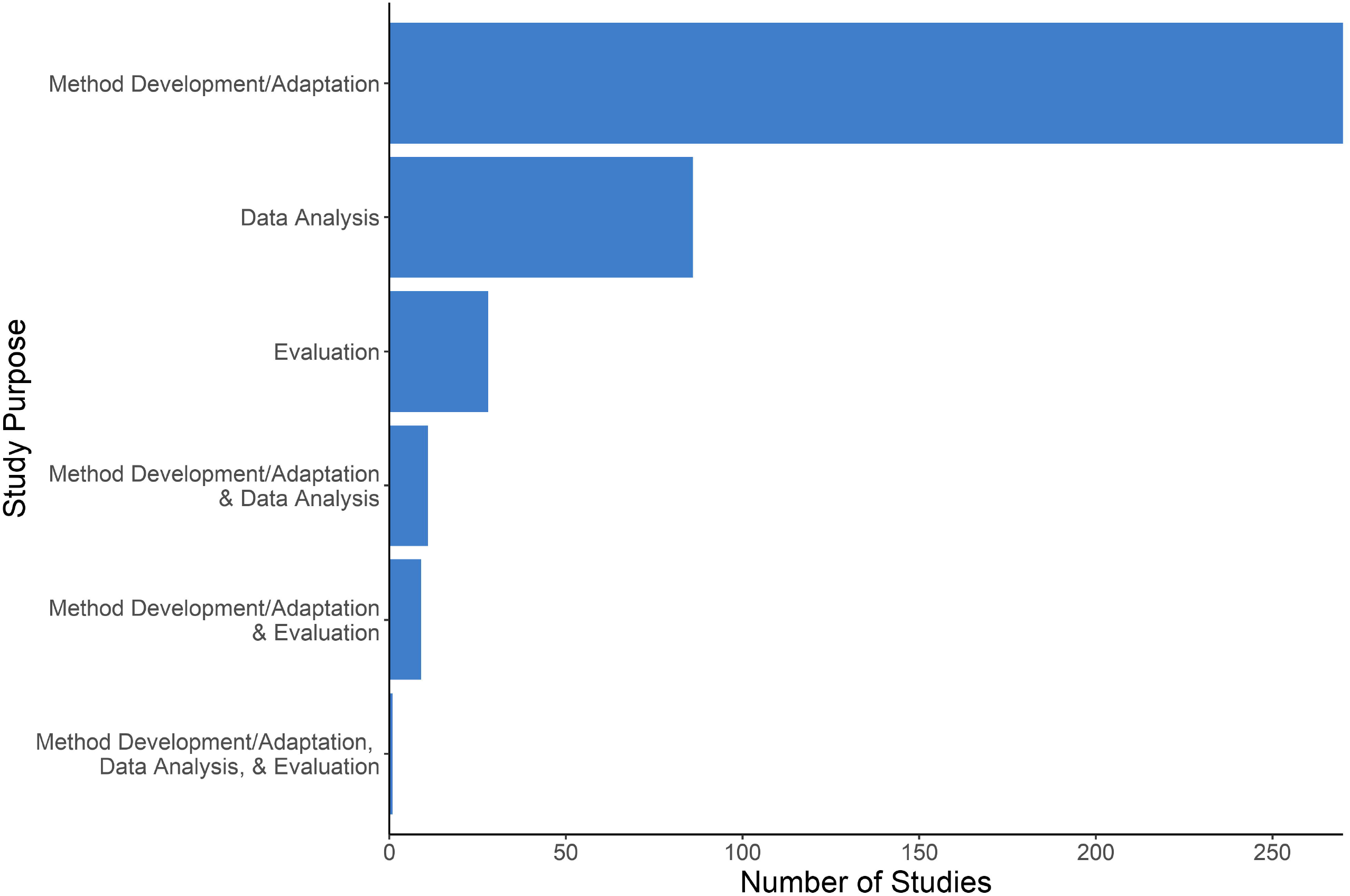
Overall Study Purpose.

### Author Appointments

We categorized author appointments into four categories: 1) Technology, Engineering, and Math (TEM) discipline, meaning an author appointed in a Department of Mathematics, Engineering, Computer Science, Informatics, and/or Statistics; 2) PC discipline, meaning an author appointed in a Department of Family Medicine, Primary Care, Community Health, and/or other analogous term; 3) Nursing discipline, and 4) Other. Authors were predominantly from TEM disciplines with 214 (52.8%) studies having at least one author with an appointment in a TEM discipline compared to 57 (14.1%) studies having at least one author with an appointment in a PC discipline. Twenty-three studies (5.7%) had a PC appointed author listed first and 27 (6.7%) had a PC appointed author listed last (senior author). These patterns did not change when unspecified or general medical appointments (i.e. any non-specialist medical appointment) were counted as PC appointments. Four studies had authors with nursing appointments. Cross tabulations between study purpose category and author appointment categories did not suggest that the types of author teams differed by study purpose. Table 1 presents a summary of the body of literature broken into PC and TEM author disciplines; Table 2S (Appendix C) contains a detailed breakdown of author appointments according to 16 categories.

### Primary Care Functions

Diagnostic decision support was by far the most common PC function present (n=148, 36.5%), followed by support for treatment decisions (n=56, 13.8%). The third most common function AI was used for was extracting information from data sources such as EMRs (n=49, 12.1%). Of studies that used AI for multiple functions, the most frequent combination was information extraction and description (n=21, 5.2%). Figure 3 summarizes the extent to which PC functions were targeted; Figure 1S (Appendix C) presents a more detailed breakdown.

**Figure 3.**
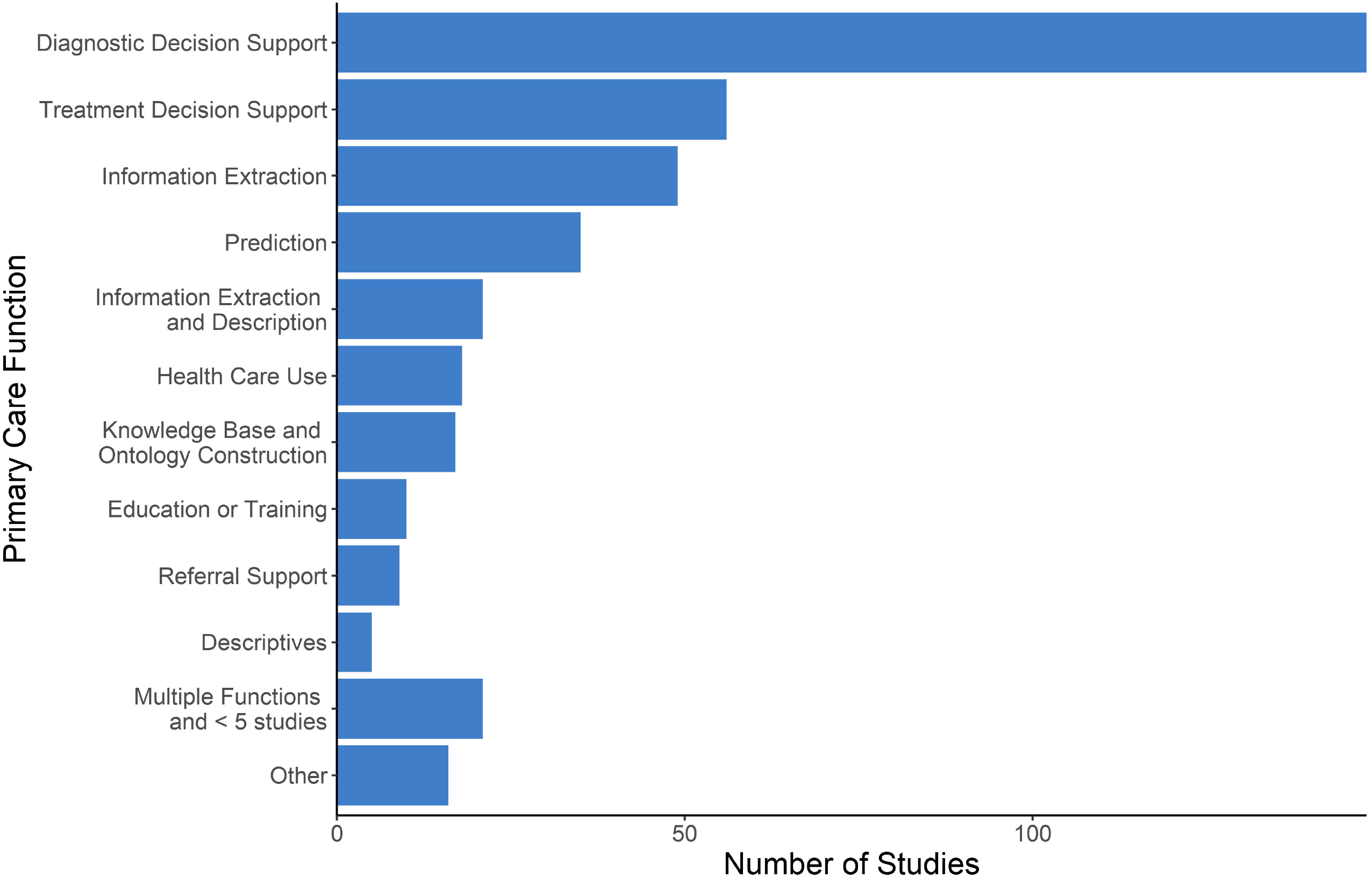
Primary Care Functions to be Support with Artifical Intelligence.

### Author Reported Target End User

The majority of studies reported physicians as a target end user of the research, either alone or in combination with other possible target end users (n=243, 60%). There appears to be no positive association between having physicians as a target end user and having at least one author with a medical appointment: the percentage of studies with at least one author with an appointment in a medical discipline of any kind was similar between studies with physician and exclusively non-physician target end-users (51.9% and 46.3%, respectively). Twenty-six (6.4 %) studies stated their research was intended for patients, 25 (6.2%) for administrative use, and 9 (2.2%) for nurses or nurse practitioners, either alone or in combination with other end users. Figure 4 shows the number of studies that included each of the target end user categories; Figure 2S (Appendix C) presents all combinations of author reported end users on a per-study basis.

**Figure 4.**
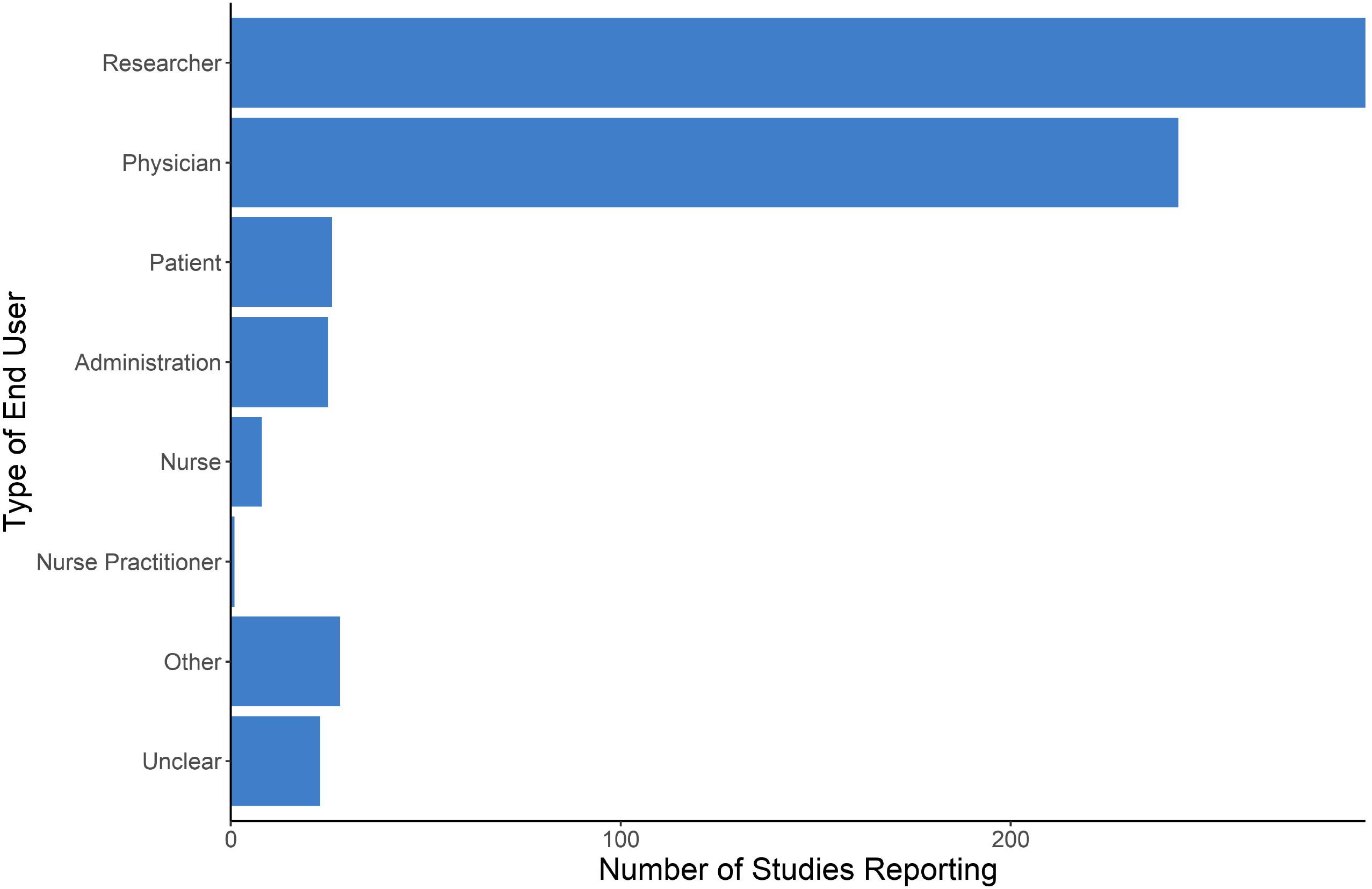
Author Reported End Users.

### Health Conditions

A large proportion of studies (n=108, 26.7%) focused on developing, using, or analyzing AI such that it would be relevant for any or for most health conditions of patients in PC settings. Of studies that targeted a particular condition, chronic physical conditions were more frequent than acute or psychiatric conditions. Target health conditions were condensed into 10 categories and are presented in Figure 5; Table 3S (Appendix C) expands the health conditions into 27 categories.

**Figure 5.**
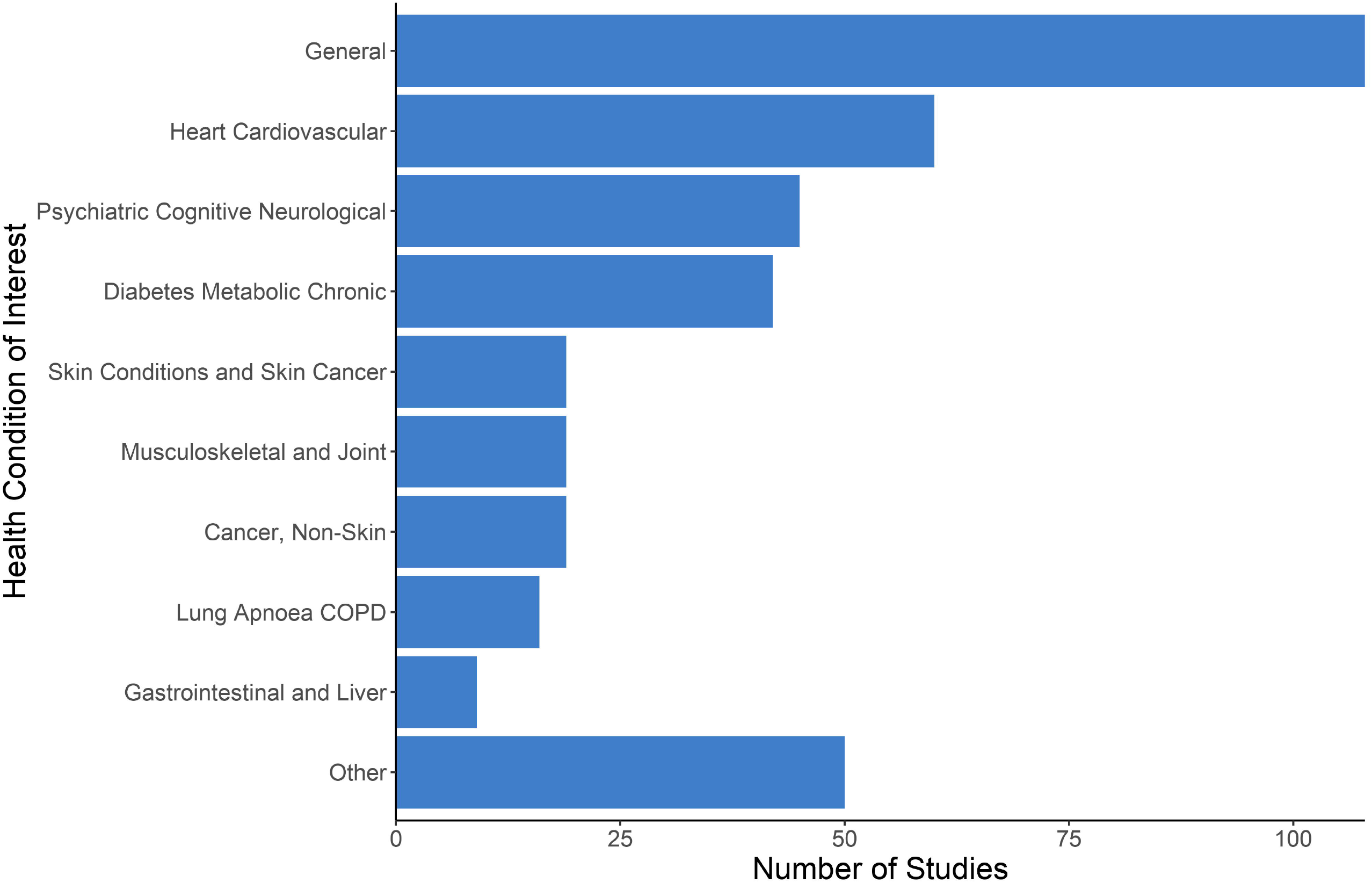
Health Conditions.

### Geographical Location

Predominantly, the location of data source(s) used in a study or the intended location of the AI implementation were OECD countries. Low- and middle-income countries were poorly represented. The majority of studies used data from a single country, with the United States alone appearing to be the most common source of data for research on AI (n=79, 19.5%). Figure 6 contains a summary of location counts and per capita rates; Table 3S (Appendix C) contains a more detailed breakdown of the counts.

**Figure 6.**
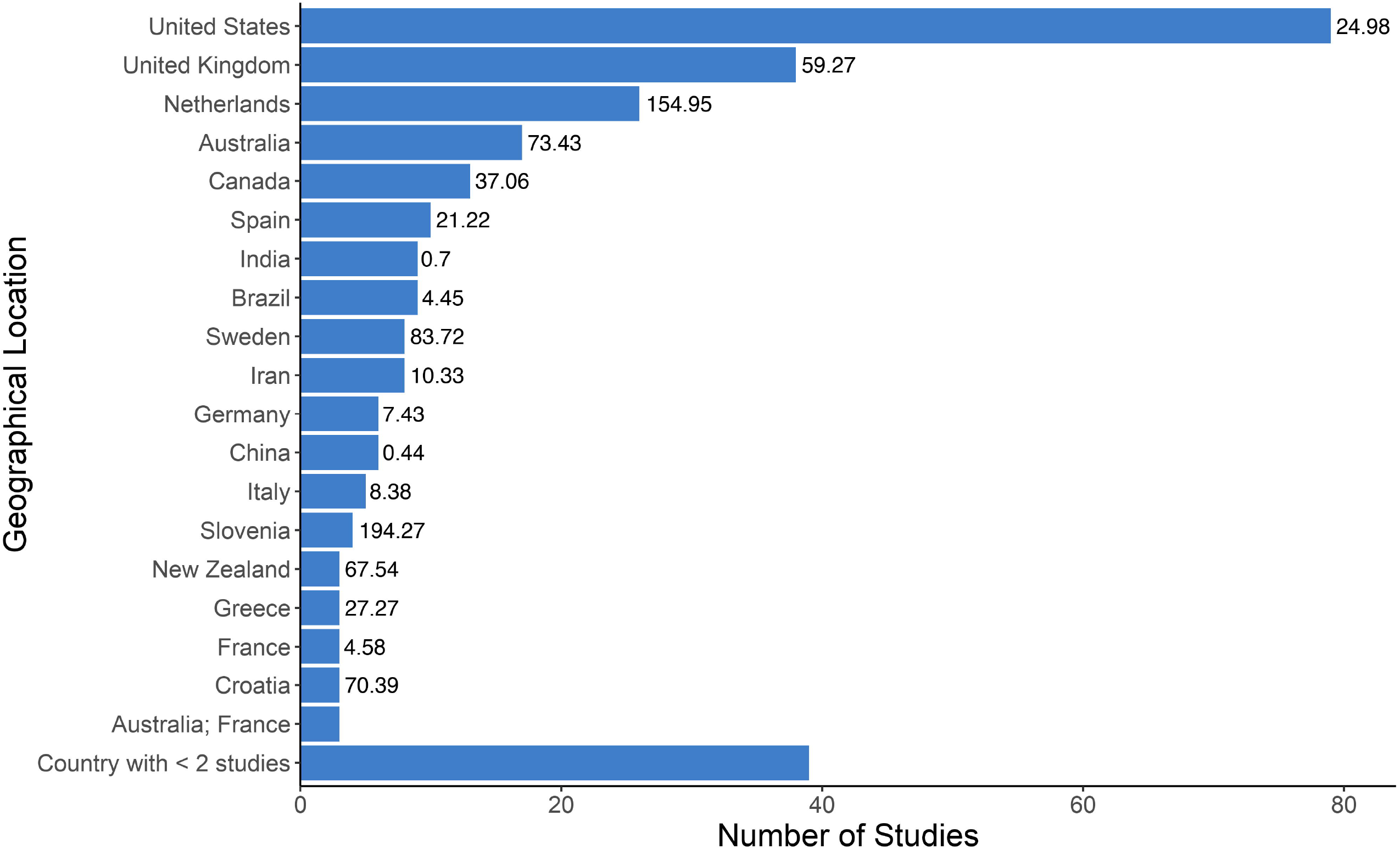
Location of Data Source or Intended Implemenation with Per Capita Rates.

### AI Subfield

The majority of studies (n=363, 89.6%) employed methods within a single subfield of AI and of these, supervised machine learning was the most common (n=162, 40.0%), followed by expert systems (n=90, 22.2%), and then natural language processing (n=35, 8.6%). There were no robotics papers. Expert systems had the earliest median year of publication (2007); data mining had the most recent (2015). Figure 7 presents frequencies and median year of publication for 10 subfields of AI used by studies captured in our literature review; all AI subfield combinations are presented in Figure 4S (Appendix C).

**Figure 7.**
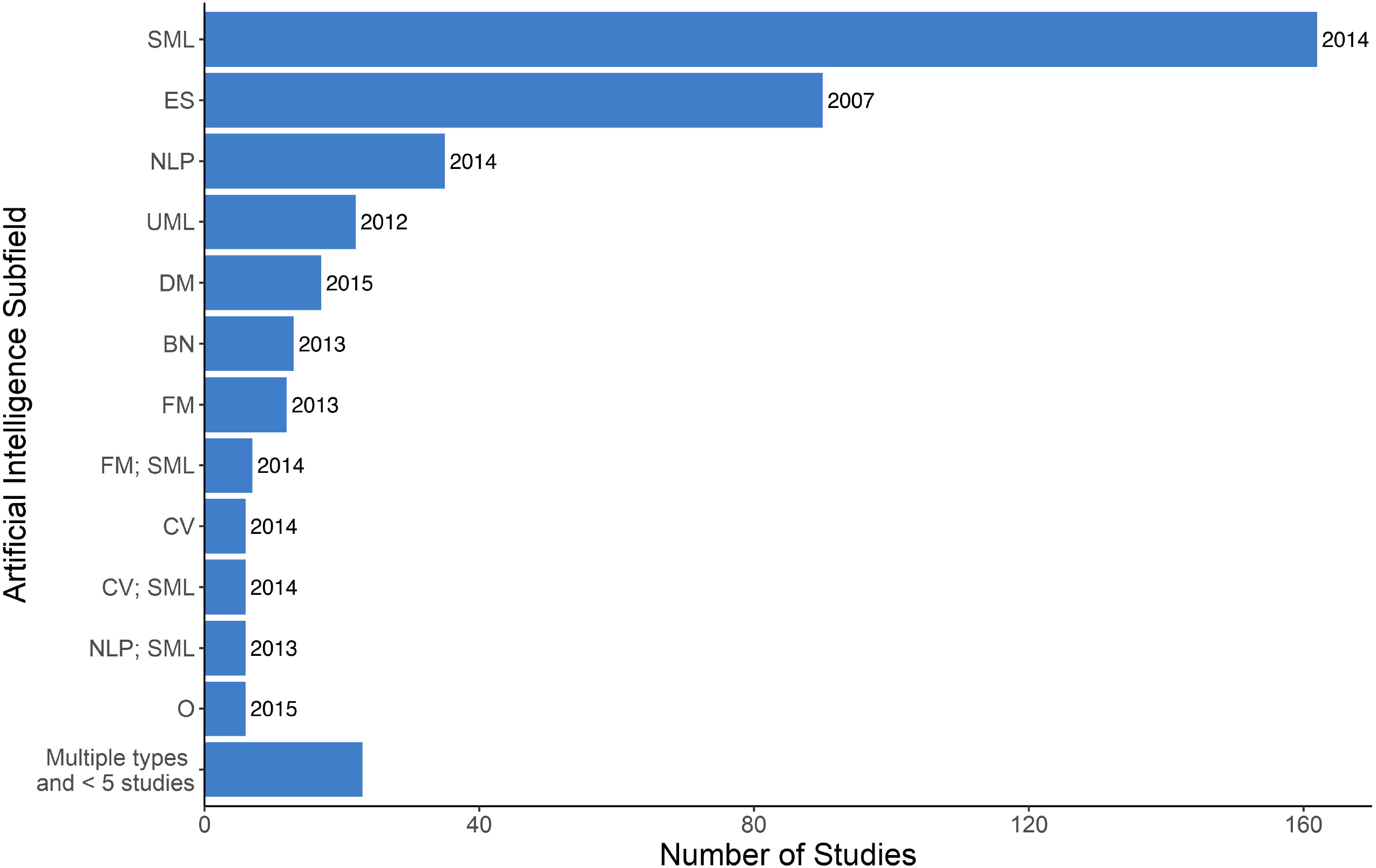
Subfields of Aritifical Intelligence with Median Year of Publication.

## DISCUSSION

### Key Findings

We located and summarized the characteristics of 405 research studies that included AI and PC. Predominant trends included 1) who: the vast majority of studies did not have any PC involvement; 2) methods: an initial focus on expert system methods shifted overtime to supervised machine learning; and 3) applications: studies most often developed AI to support diagnostic or treatment decisions, most often for chronic conditions, predominantly in higher income countries. Overall, AI and PC research is at an early stage of maturity.

The dominance of TEM-appointed authors and AI methods development research is congruent with the stage of this field. An AI-driven technology needs to be working reliably with good performance, which is accomplished through methods development research, before ‘real world’ testing, let alone widespread implementation. This is further reflected by the majority of studies having researchers as an intended end user alongside care providers, whereby further refinement seemed to be required before the AI application would be ready for use in a practice setting. For example, methods that are intended to work with many health conditions but have only been tested with a single candidate condition, or methodologically well-performing AI that needs to be incorporated into software and pilot-tested in practice settings to evaluate its impact with end-users. On the other hand, studies which focused on using AI to analyse health data may be considered distinct and are at a later stage of maturity in terms of readiness for use. These AI applications will not be used in everyday clinical practice, so while methodological performance is important, longer-term health or workflow outcomes may not need to be assessed before ‘real world’ use.

The dominant subfields of AI identified by our review mirror broader trends in AI research and make sense in terms of other characteristics of the included studies. Expert systems comprise a significant portion of the literature but are now less common (median year of publication 2007 versus 2014 for supervised machine learning), reflecting a general shift in AI research from expert systems and rule-centric AI methods to machine learning and data-centric AI methods (57). These methods are amendable to providing diagnostic and treatment recommendations as well as making predictions about future health, which supports PC activities such as primary prevention and screening. This also aligns with the focus on physicians as target eventual end users.

An underlying driver of AI research, and by extension maturation, is data availability, particularly after the shift toward data-driven machine learning methods. The United States is the single dominant country in the field, which is not surprising given its population, wealth, and research resources and output (58–61). The high standing of the United Kingdom and Netherlands despite smaller populations may be attributable to PC data availability (62,63), facilitated by high adoption rates of EMRs (64), and strong information technology academics and industries (65,66).

### Strengths and limitations

Strengths of our review include a comprehensive search strategy in peer reviewed and grey literature databases, without date restriction, with inclusive eligibility criterion, and conducted by an interdisciplinary research team. Limitations include using multiple reviewers to extract data without double coding and English language restriction. Proprietary research would not be captured by our review.

### Suggestions for future research

Our next steps include further assessing the relevance and quality of the included studies and summarizing cases of exemplary research projects. We additionally recommend a review on AI for the broader primary health care system that includes care providers beyond physicians and nurses (e.g. social workers, physiotherapists).

In terms of future research studies, there is a need for more interdisciplinary research teams with PC end user engagement. Value must be placed both on methods development contributions and on the potential impact of developed AI technologies for intended end users. Knowledge and skills from TEM disciplines will contribute towards rigorous methodology and AI performance; knowledge and skills from PC disciplines will contribute towards identifying the best PC challenges to target and assessing success in terms of care delivery and longer-term health outcomes. Inclusion of nurses, patients, and administrative staff needs to increase—identifying relevant non-physician end user activities that could be augmented by AI is an outstanding research question on its own.

For future AI methods development, we expect to see a shift towards a middle-ground between rule-centric and data-centric methods because interpretable models may better support decisions and trust in the healthcare setting. For example, explainable AI is a paradigm whereby one can understand what a model is doing or why it arrives at a particular output (67–69). Interpretability of models is additionally important from an equity lens to be able to identify and then avoid AI reproducing biases in data, which is a present concern with data-driven methods (70). It is also important to remember that AI is not always a superior solution; a recent review found no benefit overall of machine learning compared to logistic regression for clinical prediction rules (71).

Finally, availability of data determines the clinical problem areas and PC functions to which AI can be applied, and thus also a limit on where it can help. Investments in data generation, quality, and access will increase future possibilities for AI to be used to strengthen PC in the corresponding region. This includes adoption and use of EMRs (64,72).

### Conclusions

To our knowledge this is the first comprehensive, interdisciplinary summary of research on AI and PC. Two fundamental aims in the body of research on AI and PC emerged: 1) support for care provider decisions and 2) extracting meaningful information from PC data.

Overall, AI for PC is an innovation in early stages of maturity, with few tools ready for widespread implementation. Interdisciplinary research teams including front line clinicians and evaluation studies in PC settings will be crucial for advancement and success of AI for PC purposes.

## Data Availability

References to the 405 studies included in this review are available in the supplementary material. Our results were arrived at through summarizing the content of those studies.

## ACKNOWLEDGEMENTS

This research was supported by funding from the Canadian Institutes of Health Research and from a TUTOR-PHC Fellowship funded by INSPIRE-PHC.

---

**Figure 1S.**
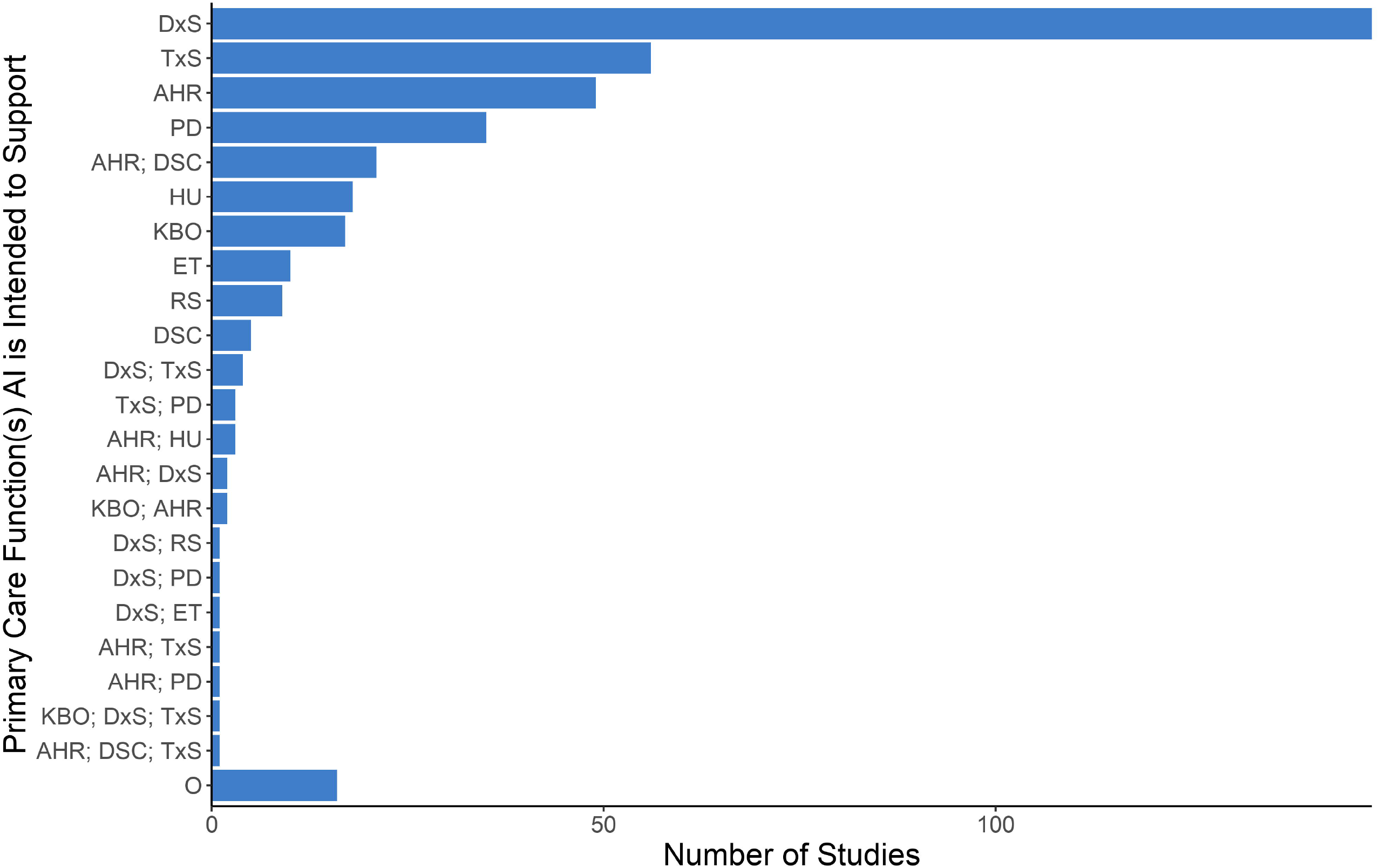
Detailed Breakdown of Primary Care Functions.

**Figure 2S:**
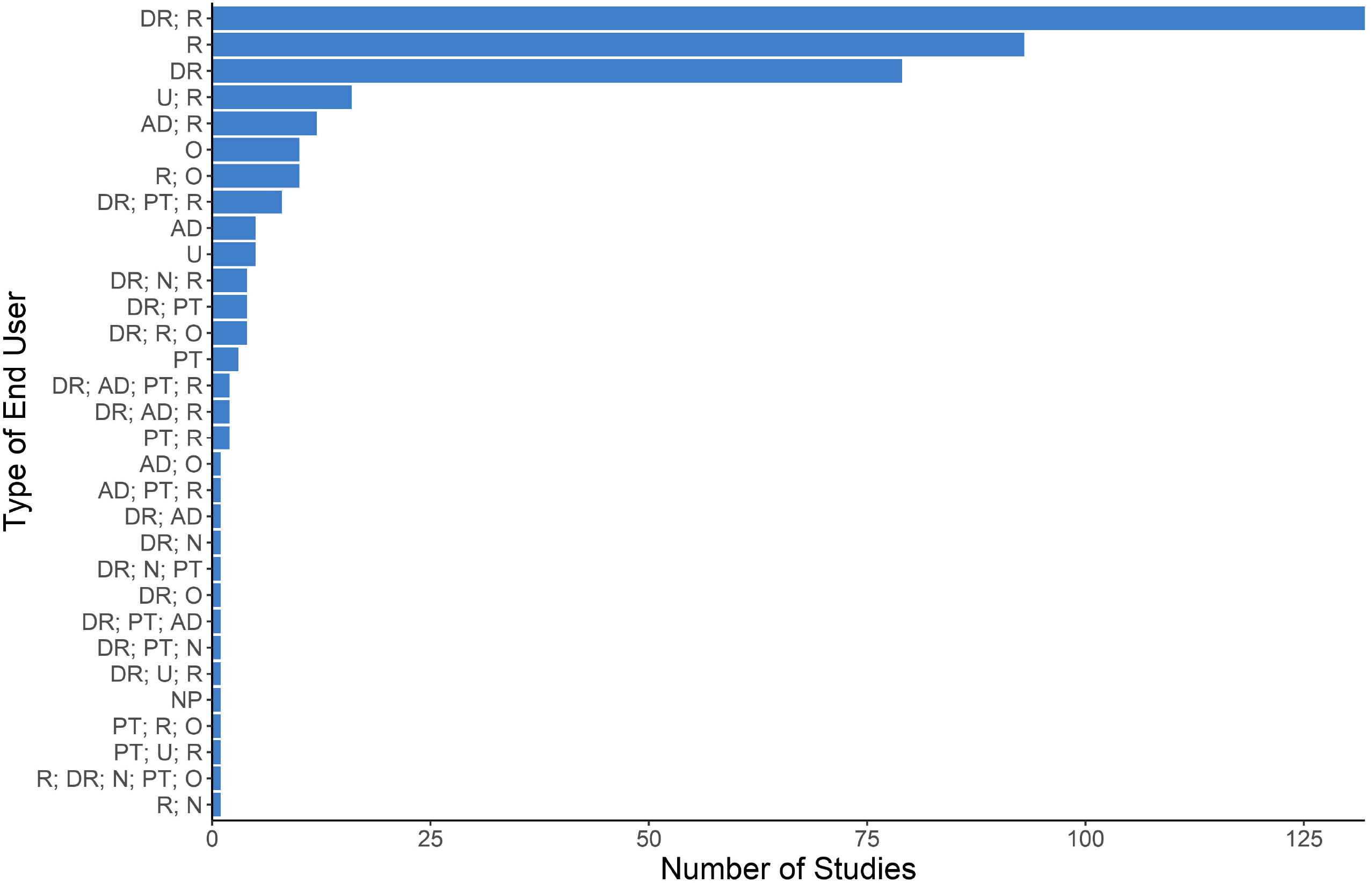
Detailed Breakdown of Author Reported Intended End Users.

**Figure 3S.**
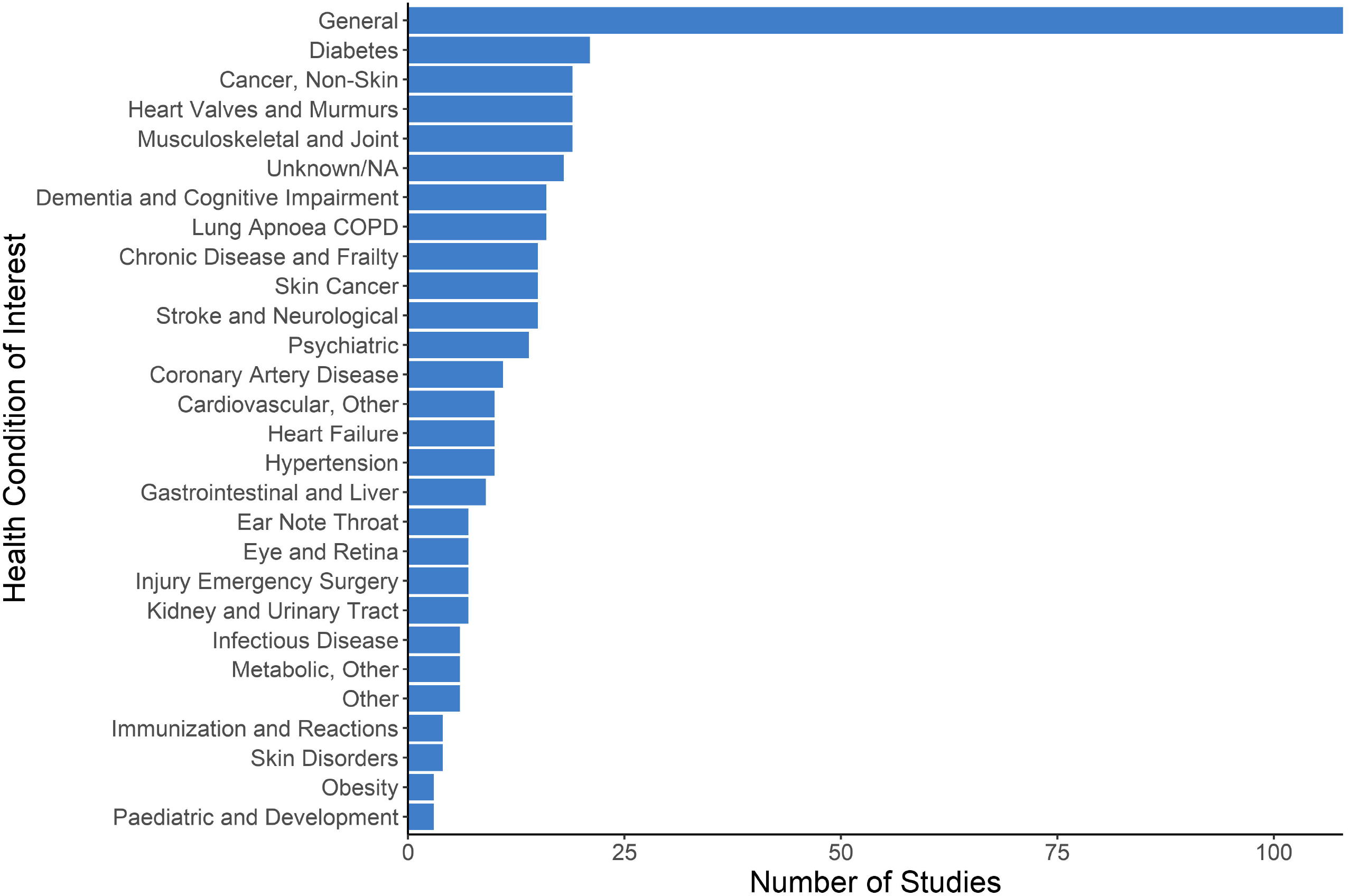
Detailed Breakdown of Health Conditions.

**Figure 4S:**
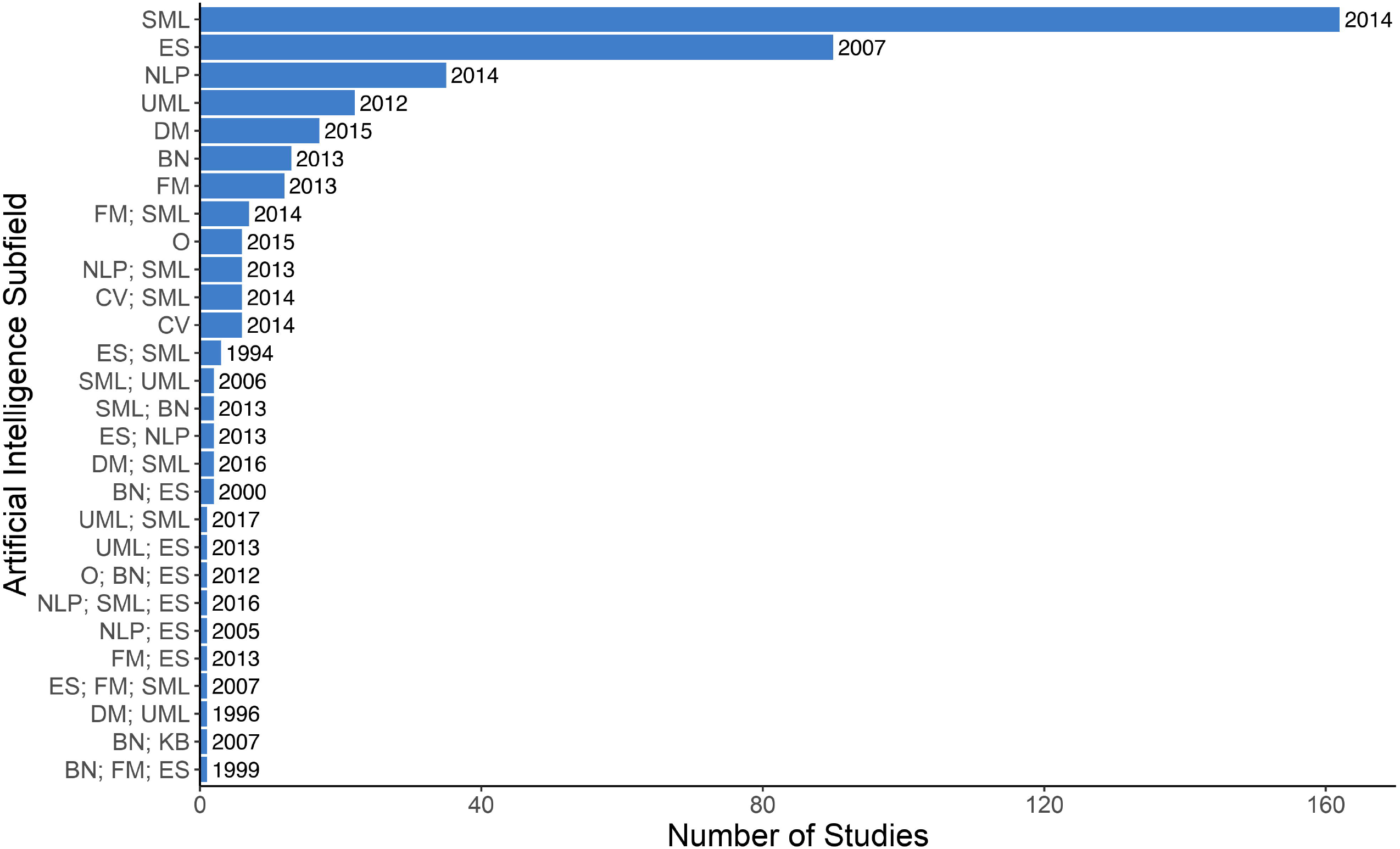
Detailed Breakdown of Aritifical Intelligence Subfields with Median Year of Publication.

